# Multilevel Factors Associated with Nonresponse to Patient-Reported Outcome Measures in Routine Radiation Oncology Care

**DOI:** 10.64898/2026.07.15.26358162

**Authors:** Jason B. Liu, Yu-Jen Chen, Maria O. Edelen, Andrea L. Pusic, Neil E. Martin, Chengbo Zeng

## Abstract

**Purpose:** Nonresponse to routinely collected patient-reported outcome measures (PROMs) threatens the representativeness of aggregated data. We characterized patient-, provider-, and clinic-level factors associated with PROMIS Global-10 nonresponse in routine radiation oncology care.

**Methods:** In this retrospective cohort study, all adults seen at five Mass General Brigham radiation oncology clinics over one year were included. The primary outcome was patient-level nonresponse, defined as never completing the portal-administered Global-10 versus completing it at least once. Using iterative mixed-effects logistic regression, we modeled patient-, provider-, and clinic-level factors.

**Results:** Among 12,214 patients, 71 providers, and five clinics, patient- and appointment-level response rates were 35.4% and 31.7%, with patient-level response ranging nearly fivefold across clinics (12.8% to 66.2%). In Model 1, male sex, lower education, not working, and recent surgery had higher odds of nonresponse, and longer time since diagnosis lower odds. After provider- and clinic-level factors were added, patient sex, education, and employment became nonsignificant, whereas recent surgery (adjusted odds ratio [aOR] 1.97) and longer time since diagnosis (aOR 0.46 for >12 months) persisted. A provider’s historical collection rate was protective but attenuated at the clinic level. There, a later program launch (aOR 0.29) and higher historical collection rate (aOR 0.79) correlated with lower nonresponse, whereas academic versus community setting did not.

**Conclusions:** Nonresponse to routinely collected PROMs is a multilevel phenomenon driven substantially by clinic-level implementation factors, not patient characteristics alone. Because response rate is only a proxy for representativeness, PROMs programs and PRO-based performance measures should prioritize representative collection over volume.

## Introduction

Patient-reported outcome measures (PROMs) are increasingly embedded in routine clinical care to foster patient-centeredness, quality and performance improvement, and value-based care transformation.^1–5^ As health systems scale PROMs collection through the electronic health record (EHR), these data are also being used to generate real-world evidence and support population-level efforts. Using PROMs data in the aggregate places a premium on data quality that extends beyond the individual patient encounter.^6^

A core threat to the utility of aggregated PROMs data is nonresponse.^7,8^ When data are systematically incomplete, analyses based on observed responses may not represent the underlying patient population, leading to biased estimates that can mask or amplify disparities.^9,10^ In routine clinical workflows, nonresponse can arise from patient-level barriers, such as language discordance or limited digital access, but it can also reflect clinic- and provider-level variation in how PROMs are operationalized, monitored, and incorporated into care.^8,11–13^ Most efforts to address nonresponse focus on raising overall response rates (aka completion rates), but this alone does not necessarily improve data representativeness, because the strategies that increase responses may not preferentially reach the patients most likely to be missing.^14,15^ Improving representativeness therefore requires first understanding where in the care delivery system nonresponse concentrates and which patients are disproportionately absent from the resulting data.

Implementation science offers an organizing framework for understanding why PROMs initiatives succeed or fail in routine care and for designing strategies to address the barriers involved.^16^ Prior work shows that the determinants of these initiatives span multiple domains and levels, from individual patients to providers and the broader care delivery system,^13,17,18^ and emphasizes implementation outcomes such as adoption, fidelity, and sustainment.^19^ Viewed through implementation science theory, missing PROMs data are less a statistical nuisance than a modifiable implementation outcome that reflects the underlying system’s design and workflow performance. Characterizing nonresponse as a multilevel phenomenon is therefore central to ensuring the representativeness of aggregated PROMs data.

Mass General Brigham (MGB) radiation oncology clinics routinely collect the Patient-Reported Outcomes Measurement Information System (PROMIS®) Global-10 across all cancer types.^20^ Every patient scheduled to see a physician or advanced practice provider (APP) is electronically administered the Global-10 before each visit, creating an opportunity to study nonresponse in a real-world setting. Drawing on established multilevel implementation science frameworks,^19,21,22^ we characterized patterns of Global-10 nonresponse across the clinic, provider, and patient levels. By identifying factors at each level, we sought to delineate whether nonresponse is shaped not only by patient characteristics but also by the providers and clinics responsible for collection. We posit that such data can strengthen the case for making data representativeness, rather than response volume alone, a core quality target for evidence generated from routinely collected PROMs.

## Methods

### Study Design, Setting, and Participants

All adults seen at five radiation oncology clinics within the MGB health system between October 2023 and September 2024 were included in this retrospective cohort study. Data were obtained from the EHR and other readily available institutional administrative data sources (e.g., credentialing information). This study was approved by the Dana-Farber/Harvard Cancer Center Institutional Review Board (24-225) and exempted from oversight.

As part of routine care, the PROMIS Global-10 version 1.0/1.1 was electronically administered to all patients via the EHR’s patient portal seven days before any visit with a provider (i.e., physician or APP).^20,23^ Patients could complete the Global-10 at any point within the 7-day window and those who did not complete the Global-10 in advance were provided electronic tablets (i.e., iPads) at the time of check-in while waiting for their appointment. Patients were required to complete all 10 Global-10 items, thus there is no item-level missingness. Per institutional policy, the Global-10 was made available in English, Spanish, Portuguese, Haitian Creole, traditional Chinese, Russian, and Arabic.^11^ Responses and scores were immediately available in the EHR for provider use.

### Definition of Nonresponse

The primary outcome for this analysis was patient-level nonresponse, defined as whether the patient completed the Global-10 one or more times during the study period (respondent) versus none (nonrespondent). Because a patient could have one or more appointments during the study period and therefore could be administered the Global-10 more than once, we also calculated appointment-level response rates as secondary outcomes.

### Multilevel Factors: Clinics, Providers, and Patients

Following published multilevel implementation frameworks, we conceptualized potential clinic, provider, and patient factors that might contribute to nonresponse using readily available data.^16,19,21,22,24^ **Supplemental Table 1** outlines the factors in detail along with corresponding implementation domains. Briefly, clinic-level factors included: clinic type, year in which PROMs collection was implemented, weekly patient volume, and weekly response rates. Provider-level factors included: sex, years since graduation from their terminal clinical degree, and role. Weekly patient volume and weekly responses rates were also calculated by provider. Patient-level demographic characteristics included: age, sex, race/ethnicity, highest level of education attainment, insurance type, employment status, preferred language, comorbidities based on the NCI version of the Charlson comorbidity index (CCI),^25^ cancer type, months since cancer diagnosis, and whether the patient had a surgical procedure within the previous six months.

### Statistical analysis

Clinic-, provider-, and patient-level characteristics were summarized using medians and interquartile range (IQRs) for continuous variables and frequencies and percentages for categorical variables. We calculated both patient- and appointment-level response rates for each clinic during the study period. Differences in clinic-, provider-, and patient-level characteristics between nonrespondents and respondents were assessed, and effect sizes (ES), as quantified with Cramér’s *V* or Cohen’s *d*. We considered ES of 0.2, 0.5, and 0.8 to reflect small, medium, and large effects.^26^

We used an iterative modeling approach to identify clinic-, provider-, and patient-level factors associated with nonresponse. To ensure model convergence and robustness, we excluded four providers with fewer than 20 patients, which we did not expect to differ substantially from those included in the analysis. First, we fit a mixed-effects logistic regression model to examine associations between patient-level characteristics and nonresponse (Model 1). Based on the magnitude of differences between nonrespondents and respondents, we included age, sex, race/ethnicity, education, insurance type, employment status, CCI, time since diagnosis, and receipt of surgery in the previous six months as fixed effects, with cancer type as a random effect. Second, patient-level fixed factors that were significant in Model 1 were retained, and provider-level fixed factors (sex, years since graduation, role, weekly collection rate, and patient volume) were added to the model, with provider as a random effect (Model 2). Finally, clinic-level fixed factors (hospital type, year of Global-10 implementation, and weekly collection rate) were further incorporated alongside significant patient- and provider-level fixed factors, with clinic as a random effect (Model 3). Results are reported as odds ratios (ORs) with 95% confidence intervals (CIs), wherein OR >1.0 represents higher odds of nonresponse. All analyses were conducted using SAS v9.4. Tests of significance were two-sided.

## Results

### Overview

Five clinics, 71 providers, and 12,214 unique patients were included (**Figure 1**). Over the 12-month study period, the Global-10 was assigned to 12,214 patients encompassing 39,510 appointments (**Table 1**). There were 4,325 patients who responded to one or more Global-10 administrations, yielding an overall patient-level response rate of 35.4%. Of the 39,510 appointments with an accompanying Global-10 assignment, 12,521 responses were obtained, yielding an appointment-level response rate of 31.7%.

**Figure 1.** Study cohort selection. FY: fiscal year

**Table 1.** PROMIS Global-10 assignments and completions at the patient and appointment levels.

| Clinics | Assignments |  | Completions |  | Response rate<br>at patient level (%)<sup>†</sup> | Response rate<br>at appointment level (%)<sup>*</sup> |
| --- | --- | --- | --- | --- | --- | --- |
|  | Patients<sup>-</sup><br>(P) | Appointments (E) | Patients<sup>-</sup><br>(p) | Appointments (e) |  |  |
| Academic sites | 6,522 | 19,449 | 2,438 | 6,531 | 37.4 | 33.6 |
| Site 1 | 4,502 | 12,613 | 1,568 | 4,105 | 34.8 | 32.5 |
| Site 2 | 2,325 | 6,836 | 962 | 2,426 | 41.4 | 35.5 |
| Community sites | 5,916 | 20,061 | 1,948 | 5,990 | 32.9 | 29.9 |
| Site 3 | 1,881 | 6,233 | 241 | 663 | 12.8 | 10.6 |
| Site 4 | 1,007 | 3,736 | 667 | 2,097 | 66.2 | 56.1 |
| Site 5 | 3,082 | 10,092 | 1,060 | 3,230 | 34.4 | 32.0 |
| Total | 12,214 | 39,510 | 4,325 | 12,521 | 35.4 | 31.7 |

\* Response rate at appointment level = $e/E * 100\%$ ; †: Response rate at patient level = $p/P * 100\%$ . □: Numbers do not sum to the total because patients were allowed to receive care across sites.

As expected, response rates varied across the clinics at both the patient and appointment levels (**Table 1**). Patient-level response rates ranged from 12.8% to 66.2% and appointment-level response rates ranged from 10.6% to 56.1%.

Two clinics were located within academic medical centers and three in community settings (**Table 2**). Clinics located at academic sites had higher response rates than community ones (patient level: 37.4% vs. 32.9%; appointment level: 33.6% vs. 22.9%). Three clinics began collecting PROMs in 2015. Weekly patient volume across clinics ranged from 23 to 161 patients. Weekly PROMs response rate ranged from 38.9% to 85.7%.

**Table 2.** Clinic characteristics (N=5).

|  | Site 1 | Site 2 | Site 3 | Site 5 | Site 6 |
| --- | --- | --- | --- | --- | --- |
| Clinic setting | Academic | Academic | Community | Community | Community |
| Weekly patient volume | 161 | 58 | 41 | 23 | 49 |
| Global-10 collection since | 2015 | 2016 | 2015 | 2017 | 2015 |
| No. of patients assigned weekly<sup>‡</sup> | 105 | 44 | 36 | 21 | 33 |
| No. of patients responded weekly | 54 | 31 | 14 | 18 | 14 |
| Weekly response rate (%)<sup>†</sup> | 51.4 | 70.5 | 38.9 | 85.7 | 42.4 |

‡: not all appointment types were accompanied with a Global-10 administration. †: Weekly response rate = No. of patients responded weekly / No. of patients assigned weekly.

Among the 71 providers, 51.4% were female and 78.9% were physicians (**Table 3**). The median time since graduation from corresponding professional schools was 12 years (IQR 8-20). Providers saw a median of 7 patients per week (IQR 4-10). The median weekly PROMs response rate was 50.0% (IQR 31.0-58.0%).

**Table 3.** Provider characteristics (N=71).

|  | N (%) |
| --- | --- |
| Sex |  |
| Female | 36 (51.4) |
| Male | 34 (48.6) |
| Years since graduation (Median, IQR) | 12 (8 - 20) |
| 1 – 8 | 23 (32.4) |
| 9 – 12 | 13 (18.3) |
| 13 – 20 | 18 (25.4) |
| ≥21 | 17 (23.9) |
| Role |  |
| Physician | 56 (78.9) |
| Advanced practice providers | 15 (21.1) |
| Weekly patient volume (Median, IQR) | 7 (4 – 10) |
| 0 – 5 | 34 (47.9) |
| 6 – 9 | 21 (29.6) |
| 10 – 12 | 8 (11.3) |
| ≥13 | 8 (11.3) |
| No. of patients assigned weekly (Median, IQR) | 5 (3 – 8) |
| No. of patients responded weekly (Median, IQR) | 2 (1 – 4) |
| Weekly response rate<sup>†</sup> (Median, IQR) | 0.50 (0.31 – 0.58) |

†: Weekly response rate = No. of patients responded weekly / No. of patients assigned weekly

**Table 4** depicts the characteristics of the 12,214 unique patients included. Overall, patients had a median age of 68 years (IQR 60-75) and slightly more were female (51.2%). Commensurate with MGB’s usual patient population, 78.8% were non-Hispanic White and 60.3% attained a college level education or higher. More than 50% of the cohort had breast or prostate cancer. 17.4% of patients had surgical procedures within the previous six months.

**Table 4.**
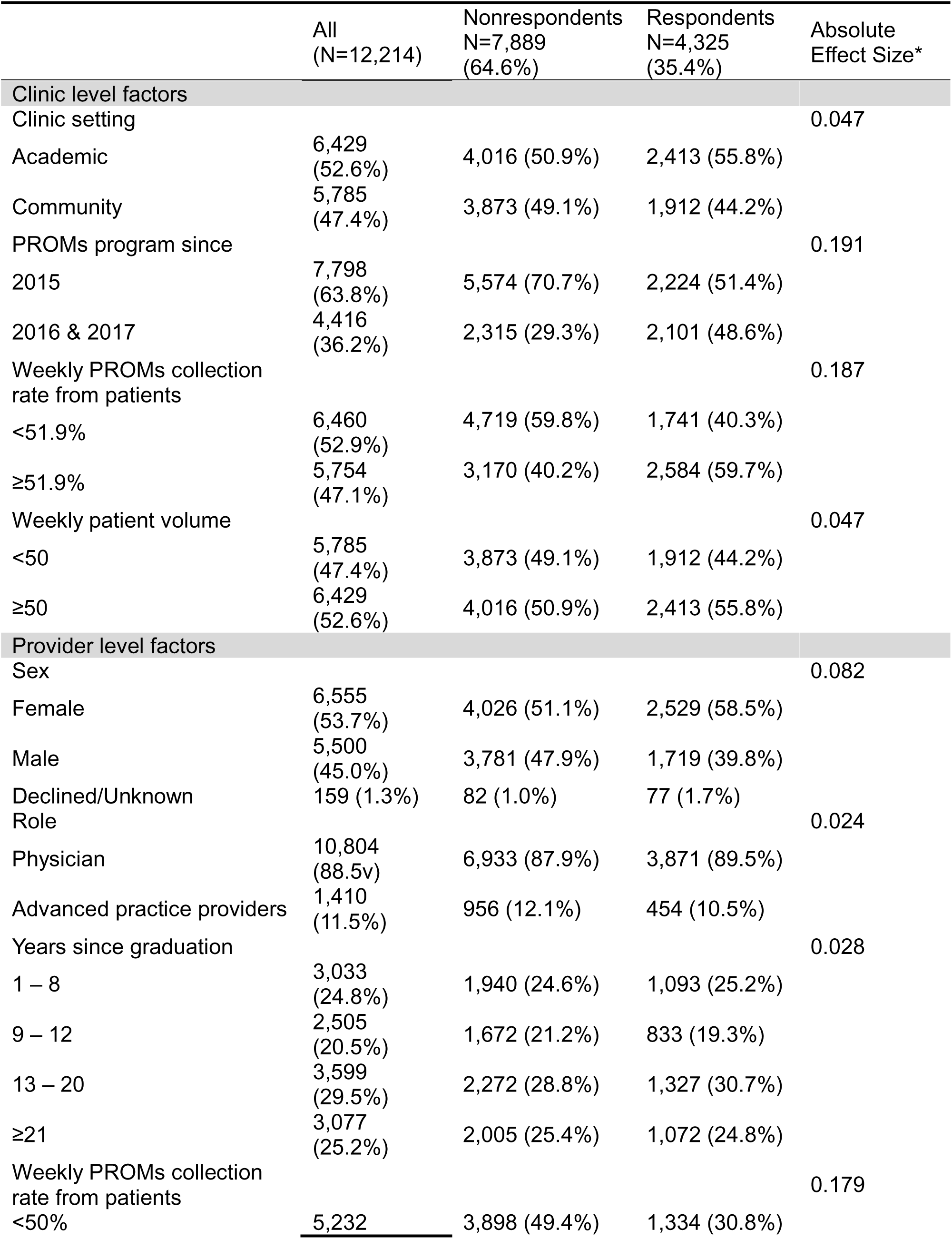
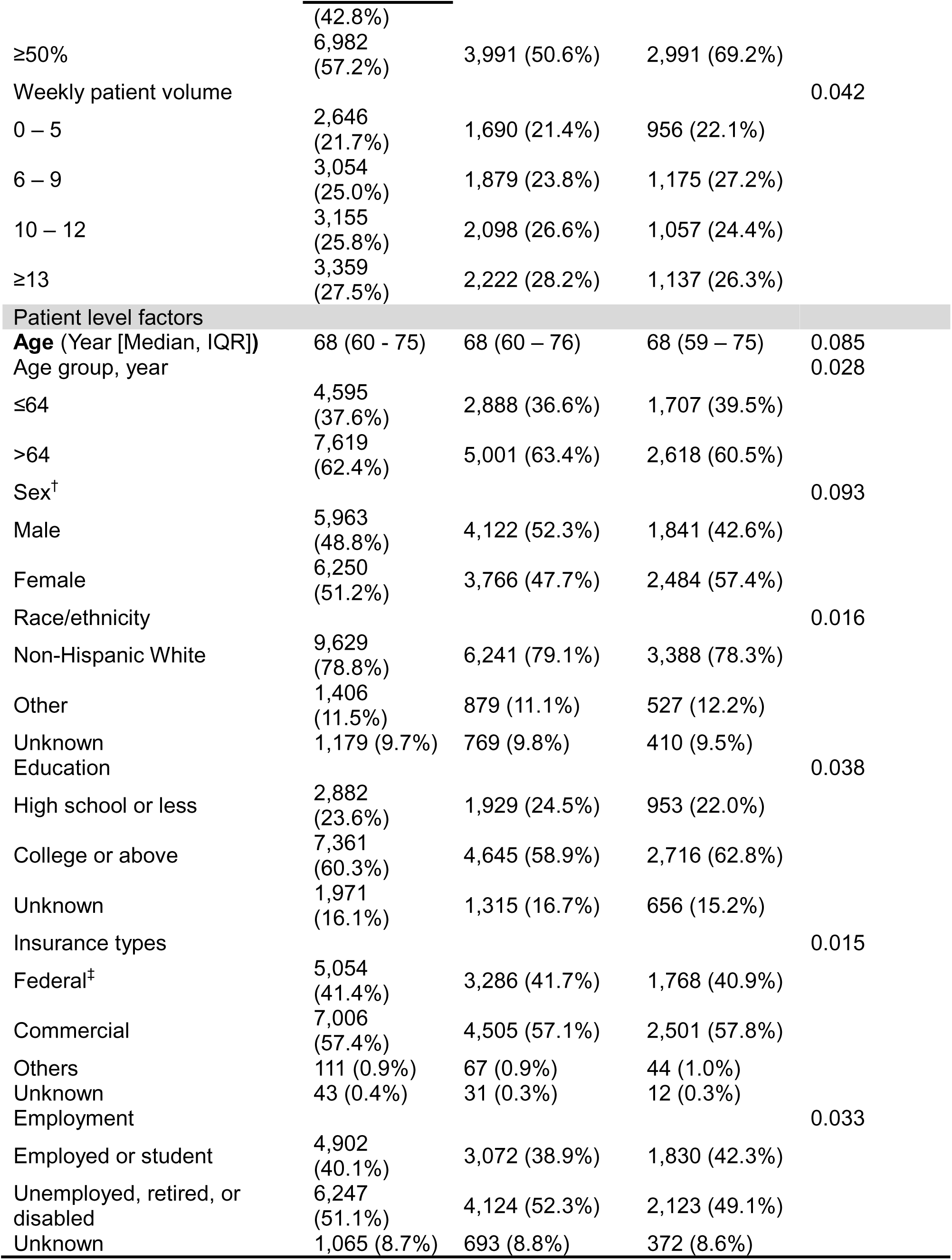
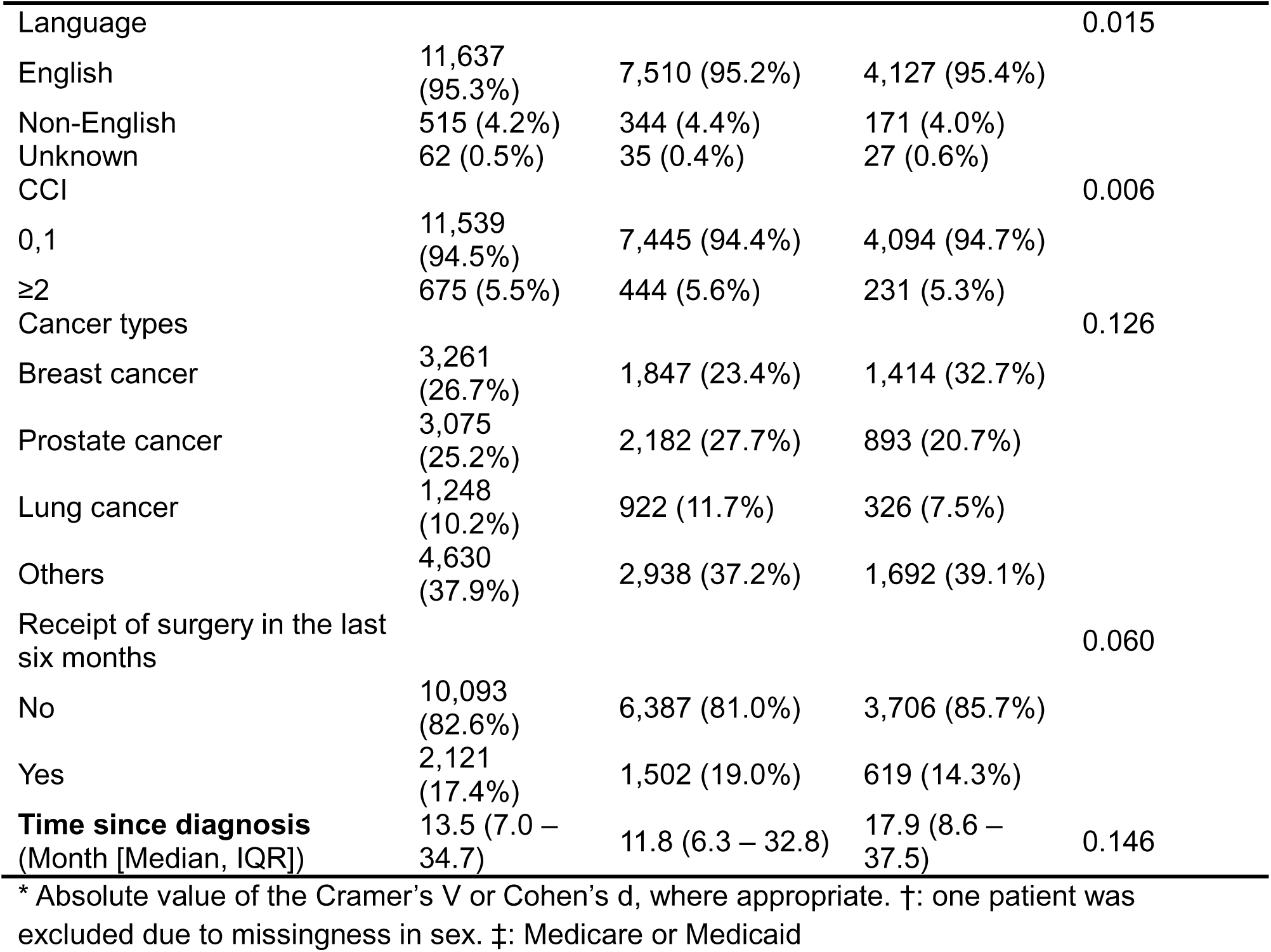
Clinic, provider, and patient characteristics by response.

### Differences between Nonrespondents and Respondents

**Table 4** depicts the differences between nonrespondents and respondents at the three levels. There was a higher proportion of respondents in clinics that began collecting PROMs in 2016 or later as compared to those in 2015 (48.6% vs. 29.3%; ES=0.191). Clinics with a weekly response rate of at least 51.9% in the year prior to the study period (the median rate across the five clinics) had a higher proportion of respondents (59.7% vs. 40.2%; ES=0.187).

Differences in provider characteristics between nonrespondents and respondents were generally small, with the exception that more responses were obtained from providers whose weekly response rate was at least 50.0% in the year preceding the study period (69.2% vs. 50.6%; ES=0.179).

Among the 12,214 patients, larger differences between nonrespondents and respondents were observed for sex, cancer type, surgery in the previous six months, and time since diagnosis. For example, we observed more female respondents than male ones (57.4% vs. 42.6%; ES=0.093). Compared with nonrespondents, respondents included a higher proportion of patients with breast cancer (32.7% vs. 23.4%) and lower proportions of patients with prostate (20.7% vs. 27.7%) and lung cancer (7.5% vs. 11.7%; ES=0.126). Among respondents, there was a lower proportion of patients who had surgery in the previous six months (14.3% vs. 19.0%; ES=0.060). In addition, respondents had a longer duration since cancer diagnosis than nonrespondents (17.9 vs. 11.8 months; ES=0.146).

### Iterative Modeling Results: Clinic-, Provider-, and Patient-level Factors Associated with Nonresponse

Iterative modeling began at the patient level and built upwards (**Table 5**). Results from Model 1, which included only patient-level factors, showed that patients who were male (adjusted odds ratio [aOR] 1.20; 95% CI 1.07–1.34), were unemployed, retired, or disabled (aOR 1.12; 95% CI 1.02–1.23), attained a high school education or less (aOR 1.14; 95% CI 1.04–1.26), or had surgery in the previous six months (aOR 1.54; 95% CI 1.37–1.71) had higher odds of being nonrespondents compared with their respective counterparts. In contrast, patients who were diagnosed between 6-12 months ago (aOR 0.80; 95% CI, 0.71–0.91) or more than 12 months ago (aOR 0.50; 95% CI, 0.45–0.56) had lower odds of being nonrespondents than those diagnosed within six months.

**Table 5.** Multilevel factors associated with nonresponse. Iterative multivariable modeling results.

| **Variables** | **Model 1** | **Model 2** | **Model 3** |
| --- | --- | --- | --- |
| ***Clinic level*** |  |  |  |
| Community hospitals |  |  | 0.81 (0.31 – 2.14) |
| Academic hospitals |  |  | Reference |
| PROM collection since 2016 |  |  | 0.29 (0.12 – 0.70)* |
| 2015 |  |  | Reference |
| Weekly collection rate ≥ 51.9% |  |  | 0.79 (0.68 – 0.92)* |
| < 51.9 % |  |  | Reference |
| ***Provider level*** |  |  |  |
| Male |  | 1.09 (0.76 – 1.56) | - |
| Unknown sex |  | 0.69 (0.18 – 2.67) | - |
| Female |  | Reference |  |
| Advanced practice providers |  | 1.30 (0.81 – 2.10) | - |
| Physicians |  | Reference |  |
| 9 – 12 years since graduation |  | 1.10 (0.84 – 1.44) | - |
| 13 – 20 years since graduation |  | 1.23 (0.89 – 1.70) | - |
| Longer than 21 years since graduation |  | 1.11 (0.77 – 1.61) | - |
| 1 – 8 years |  | Reference |  |
| Weekly collection rate ≥ 50% |  | 0.75 (0.63 – 0.89)* | 0.89 (0.76 – 1.06) |
| < 50 % |  | Reference | Reference |
| 6 – 9 patients per week |  | 0.80 (0.63 – 1.01) | - |
| 10 – 12 patients per week |  | 0.84 (0.63 – 1.11) | - |
| More than 13 patients per week |  | 0.89 (0.62 – 1.27) | - |
| 0 – 5 patients |  | Reference |  |
| ***Patient level*** |  |  |  |
| Age > 64 years | 0.97 (0.87 – 1.07) | - | - |
| ≤ 64 years | Reference |  |  |
| Male | 1.20 (1.07 – 1.34)* | 1.05 (0.92 – 1.19) | - |
| Female | Reference | Reference |  |
| Racial/ethnic minority | 0.90 (0.79 – 1.01) | - | - |
| Unknown race/ethnicity | 0.99 (0.85 – 1.15) | - | - |
| Non-Hispanic White | Reference |  |  |
| High school or less | 1.14 (1.04 – 1.26)* | 1.10 (0.99 – 1.21) | - |
| Unknown education | 1.20 (1.06 – 1.36)* | 1.09 (0.95 – 1.24) | - |
| College or above | Reference | Reference |  |
| Federal insurance | 0.98 (0.90 – 1.08) | - | - |
| Other insurance | 0.85 (0.57 – 1.26) | - | - |
| Unknown insurance | 1.55 (0.76 – 3.15) | - | - |
| Commercial insurance | Reference |  |  |
| Unemployed, retired, or disabled | 1.12 (1.02 – 1.23)* | 1.07 (0.98 – 1.17) | - |
| Unknown employment | 1.00 (0.84 – 1.19) | 0.92 (0.77 – 1.09) | - |
| Employed, student | Reference | Reference |  |
| CCI ≥ 2 | 0.96 (0.81 – 1.14) | - | - |
| CCI ≤ 1 | Reference |  |  |
| Received surgery in the last six months | 1.54 (1.37 – 1.71)* | 1.68 (1.49 – 1.89)* | 1.87 (1.65 – 2.11)* |
| No | Reference | Reference | Reference |
| 6 – 12 months since diagnosis | 0.80 (0.71 – 0.91)* | 0.83 (0.73 – 0.94)* | 0.83 (0.73 – 0.94)* |
| Longer than 12 months since diagnosis | 0.50 (0.45 – 0.56)* | 0.47 (0.42 – 0.53)* | 0.46 (0.41 – 0.52)* |
| 0 – 6 months | Reference | Reference | Reference |
| **Model fit** |  |  |  |
| AIC | 14782.89 | 13938.50 | 13886.71 |
| BIC | 14772.45 | 13896.50 | 13864.71 |

\*: p<0.05

Results from Model 2 showed that after adding provider-level factors, patient-level sex, education, and employment status were no longer significantly associated with nonresponse (**Table 5**). Having had surgery in the previous six months (aOR 1.68, 95% CI 1.49-1.89) and a longer duration of time since diagnosis (e.g., >12 months vs. 0-6 months; aOR 0.47, 95% CI 0.42-0.53) remained significantly associated with nonresponse. Notably, among the provider-level factors examined, only weekly collection rate was significantly associated with, and protective against, nonresponse (aOR 0.75; 95% CI 0.63–0.89).

After adding clinic-level factors (Model 3), the association between provider-level weekly collection rate and nonresponse was attenuated and no longer statistically significant (aOR 0.89, 95% CI 0.76-1.06; Table 5). Having had surgery in the previous six months (aOR 1.97, 95% CI 1.65-2.11) and a longer duration of time since diagnosis (e.g., >12 months vs. 0-6 months; aOR 0.46, 95% CI 0.41-0.52) remained significantly associated with nonresponse. Clinic type was not significantly associated with nonresponse (aOR 0.81; 95% CI 0.31–2.14). Consistent with the bivariate analyses, patients from clinics that began PROMs collection in or after 2016 had lower odds of nonresponse compared with those from clinics that began collecting PROMs in 2015 (aOR 0.29; 95% CI 0.12–0.70). In addition, clinic-level weekly collection rate was significantly associated with, and protective against, nonresponse (aOR 0.79; 95% CI 0.68–0.92).

## Discussion

In this study, among 12,214 patients assigned the PROMIS Global-10 across five radiation oncology clinics over a year, about one in three responded at least once (35.4%). Patient-level response varied nearly fivefold across clinics, from 12.8% to 66.2%. When nonresponse was modeled iteratively from the patient level upward, several patient characteristics that appeared influential in isolation, including male sex, lower educational attainment, and being unemployed, retired, or disabled, lost statistical significance once provider- and clinic-level factors were introduced. Two patient-level signals, however, persisted across every model. Recent surgery within six months was associated with higher odds of nonresponse (aOR 1.97), and longer time since diagnosis with lower odds (aOR 0.46 for >12 months versus ≤6 months). At higher levels, a provider’s historical collection rate was protective until clinic-level factors were added, after which clinic attributes, namely an earlier collection launch year and a higher historical collection rate, emerged as the dominant correlates of nonresponse. Academic versus community setting was not.

The central implication of these results is that nonresponse to routinely collected PROMs is not primarily a patient-level phenomenon but a property of the measurement system as a whole. The attenuation of patient-level associations once provider and clinic factors were modeled supports this notion. Male sex, for example, was significantly associated with nonresponse only when patient characteristics were considered but became nonsignificant once provider factors entered the model, and education and employment followed the same pattern. Much of what might be read as patient unwillingness or inability to respond might be absorbed by where a patient is seen and by how consistently that setting collects the PROMs. Framed through implementation science, missing PROMs data behave less like a fixed patient trait and more like a modifiable implementation outcome, an expression of adoption, fidelity, and sustainment at the clinic and provider levels rather than only a simple accumulation of individual barriers.^16,21,22^ Prior efforts to increase response rates have often emphasized the mode and delivery of administration, yet the evidence that mode alone closes the gap is mixed.^8,27,28^ Prior studies generally find electronic and paper administration to be comparable, even as the specific channel and contact strategy can shift completion at the margins. Mode is thus one lever among many. Consistent with earlier analyses showing that nonresponse to routinely collected PROMs tracks both patient and organizational characteristics, our findings argue that durable gains will come from attending to the full multilevel system in which the PROMs are embedded.^12,13^

Two patient-level findings nonetheless were robust to adjustment and warrant clinical interpretation. Patients who had undergone surgery in the prior six months and those earlier in their disease course were more often nonrespondents. The association with recent surgery was not only stable but strengthened as higher-level factors were added, and respondents had a longer median time since diagnosis than nonrespondents. A potential explanation is that completion may be lowest during the most acute and disruptive phases of cancer care, around diagnosis and the perioperative period, and rises as patients settle into other treatment phases with different touchpoint frequencies. Our data cannot confirm this directly. Because nonresponse was defined at the patient level, as completing the Global-10 at least once versus never, and we did not capture where in a patient’s treatment course a given response fell, we could not observe whether completion rose within individual patients as their care progressed. It is also conceivable that patients were prompted to respond to other PROMs or repeated Global-10 administrations by other members of the cancer care team during these time periods, which may have decreased their willingness to respond during the observed study period.^29^ If completion does follow the care trajectory in this way, nonresponse is partly a function of that trajectory itself, with implications for when and how the PROM is administered, not simply whether it is.

None of the provider characteristics we examined, including sex, role, years since graduation, and weekly patient volume, was associated with nonresponse. Only the provider’s own historical collection rate was protective, and it lost significance once clinic factors were added. This attenuation is itself informative. Rather than indicating that providers do not matter, it suggests that provider-level collection could be embedded in the norms, leadership, and resourcing of the clinic in which a provider works, consistent with evidence that staff engagement with PROMs tracks the organizational support surrounding it.^13,23^ The determinants that implementation science most often emphasizes at this level, including provider familiarity, perceived value, and a sense of ownership over PROMs, are attitudinal rather than concrete provider characteristics, and our administrative data could not capture them.^24^ Disentangling provider conviction from clinic culture is a question these data leave open and that future study using robust qualitative inquiry is positioned to answer.

At the clinic level, the two strongest correlates, year of program launch and historical collection rate, suggest program maturity and sustainment as plausible determinants. That academic and community settings did not differ after adjustment suggests setting type is a poor proxy for the underlying determinants of nonresponse.^20^ Indeed, the two community clinics had response rates at the opposite extremes of the entire cohort, a more than fivefold gap between sites of the same type. What distinguishes high-from low-collecting clinics is how robustly and consistently the program has been operationalized over time. The finding that more recently launched clinics had lower odds of nonresponse than the earliest adopters supports this. It may reflect cohort or workflow differences, but it is also consistent with the possibility that collection effort decays as programs age, such as the loss of a champion or workforce turnover. Either way, this temporal phenomenon reflects an erosion of sustainment that any single cross-sectional response rate would obscure. These patterns reinforce the need to better identify determinants of sustainability and sustainment.^19,30^

Finally, these findings bear on how the quality of aggregated PROMs data should be judged. The quantity of interest is the representativeness of the responding sample, for which the response rate is only a proxy. Survey methodology has long established that nonresponse bias depends on two terms: the response rate and the difference between respondents and nonrespondents on the quantity of interest. Expectedly, respondents and nonrespondents differed in this study using routinely collected EHR data. For instance, breast cancer patients were overrepresented among respondents and prostate and lung cancer patients underrepresented, a skew the overall response rate of 35.4% gave no indication of. To the extent that quality of life measured by the Global-10 differs across these cancer types, as is clinically expected, a group-level score drawn from respondents would reflect those who responded more than the full assigned population, and raising completion without regard to who is added would not necessarily close that gap. This is the same selection problem encountered in retrospective cohort and other observational designs, where inclusion is conditioned on a non-random mechanism even though no “response rate” is reported.^31,32^ If PROMs data are to be considered equivalent to other clinical data (e.g., laboratory values, vital signs, etc.), then concerns about response rates should eventually vanish. For now, PROMs research actually offers an advantage to the selection bias encountered in these research designs. Because every assigned patient is known, the sampling frame is observed rather than inferred, which allows PROMs researchers to characterize how respondents differed from nonrespondents directly and makes the resulting bias not only describable but, in principle, addressable. When random sampling is impractical or impossible, the survey literature has formalized this reorientation into practical tools, including representativeness indicators (R-indicators) and adaptive and responsive designs that steer collection toward underrepresented groups rather than raw completion.^33,34^

This study has limitations. First, many of the factors examined are indirect or proxy measures of the implementation domains they are meant to represent.^24^ Historical collection rate, for instance, stands in for fidelity and sustainment but does not directly capture workflow, staffing, or local collection behavior. We used the measures available in the EHR and administrative data because direct implementation metrics were not prospectively captured, a limitation that is itself instructive. It illustrates why PROMs programs should be planned from the outset to track implementation outcomes such as adoption, fidelity, and sustainment, both to manage collection and to make representativeness assessable. Second, the study included five clinics within a single health system over one fiscal year, which limits generalizability and precludes characterization of temporal trends within the study window. With only five clinics, precision and causal inference at the clinic level are constrained. Third, the analysis relied on retrospective EHR and administrative data, so unmeasured determinants of nonresponse, such as patient digital access, portal enrollment, and health literacy, could not be captured directly, and residual confounding is present.^11^ Similarly, we could not account for individual clinician practice patterns, specifically whether an APP was seeing patients in tandem with a physician or independently at separate clinic visits. Fourth, we characterized the selection mechanism, that is, who completed the PROM, but not its downstream effect on aggregated Global-10 scores as this was not the study intent. Finally, excluding providers with fewer than 20 patients to ensure model stability may have removed lower-volume providers whose collection patterns differ.

Taken together, these findings argue for a shift in how health systems frame missing PROMs data, away from maximizing the response rate and toward the representativeness of aggregated data as the operative quality target. Because the determinants of nonresponse span patient, provider, and clinic levels, interventions confined to any one level, such as patient-facing nudges (e.g., text message reminders) or changes in administration mode, are unlikely to suffice.^27,35^ The stakes are concrete in the US, where payers increasingly operationalize the response rate as a compliance requirement.^2,36,37^ The most contemporary example is the Centers for Medicare & Medicaid Services’ risk-standardized PRO-based performance measure (PRO-PM) for elective primary total hip or knee arthroplasty, which requires matched preoperative and 12-month postoperative PROs for at least 50% of eligible patients, with public reporting and payment consequences for hospitals that fall short. Such a threshold certifies the volume of data collected, not whether the responding sample resembles the eligible population, so a hospital can satisfy the requirement while computing its performance estimate on a systematically selected subsample.^14,15^ The supporting PROMs are well validated, so the concern is not whether they capture pain and function but who completes them. The more than fivefold variation in clinic-level response observed in this study shows how much an aggregate completion figure can conceal. The resulting risk-standardized improvement rate may then reflect who responded as much as how well the hospital performed, and could obscure rather than surface disparities. While this specific arthroplasty PRO-PM does account for nonresponse bias, tying payment to a response rate threshold may unintentionally reward easy-to-reach responses over balanced representation.^37^ PRO-PM design, and routine PROMs programs more broadly, would therefore benefit from explicit representativeness safeguards, including monitoring of representativeness indicators and subgroup completion, weighting or adjustment for nonresponse, or targeting of collection toward underrepresented patients, alongside continuous tracking of sustainment rather than volume alone.^12,15,38,39^ Our multilevel results suggest returning subgroup completion rates to individual clinics and providers rather than issuing uniform systemwide reminders, and concentrating assistance on the patients and points in the care trajectory where nonresponse is greatest, including staff-supported or in-language support for groups that monitoring identifies as underrepresented and additional attempts during the perioperative period.

In conclusion, as routinely collected PROMs are increasingly aggregated to generate real-world evidence, drive quality improvement, and underpin value-based and outcomes-based policy, ensuring that these aggregated data represent the patients they are meant to describe is foundational to their value. Achieving that representativeness, however, requires optimizing the PROM measurement system as a whole, across all of its levels.

## Data Availability

Individual patient data will not be shared.

## Acknowledgements

This work was supported by the National Cancer Institute (NCI) under R03CA292987. Unrelated to this work, MOE, ALP, and CZ are supported by NCI R01CA280619. The authors are deeply grateful for the tireless efforts of the entire MGB Radiation Oncology staff who stood up these efforts, especially Ms. Colleen M. Whitehouse, RN.

## Statements and Declarations

### Author Contributions

Jason B. Liu contributed to conceptualization, methodology, formal analysis, and funding acquisition, and wrote the original draft. Yu-Jen Chen contributed to data curation and formal analysis. Maria O. Edelen contributed to conceptualization and methodology. Andrea L. Pusic contributed to conceptualization, provided resources, and supervised the work. Neil E. Martin contributed to conceptualization, provided resources, and supervised and administered the project. Chengbo Zeng contributed to conceptualization, methodology, and formal analysis, supervised and administered the project, and contributed to funding acquisition. All authors reviewed and edited the manuscript and approved the final version.

---

**Supplemental Table 1.** Multilevel factors analyzed in this study.

| Multiple Level | CFIR Domain | Variable | Details |
| --- | --- | --- | --- |
| Clinic (Macro and meso) | Outer Setting Domain | Type | Academic or community setting |
|  | Inner Setting Domain (e.g., structural characteristics, implementation climate<sup>‡</sup>) | Weekly Patient Volume | Number of all patient appointments of any type per week by clinic |
|  |  | PROMs Collection Since | Year the clinic implemented PROMs collection in routine care |
|  |  | Number of Patients Assigned Weekly | Number of patient appointments with accompanying PROMs per week by clinic |
|  |  | Number of Patients Responding Weekly | Number of patient appointments with responses to accompanying PROMs per week by clinic |
|  |  | Weekly Clinic Response Rate | Number of Patients Responding Weekly divided by the Number of Patients Assigned Weekly by clinic |
| Provider (Micro) | Individuals Domain (e.g., roles, other personal attributes<sup>‡</sup>, innovation deliverers, and capability, motivation) | Sex | Provider sex; female or male |
|  |  | Years Since Graduation | Number of years since graduation from terminal clinical degree (e.g., MD for physicians) |
|  |  | Type | Physician or advanced practice provider (e.g., physician assistant, nurse practitioner) |
|  |  | Weekly Patient Volume | Number of all patient appointments of any type per week per provider |
|  |  | Number of Patients Assigned Weekly | Number of patient appointments with accompanying PROMs per week by provider |
|  |  | Number of Patients Responding Weekly | Number of patient appointments with responses to accompanying PROMs per week by provider |
|  |  | Weekly Provider Response Rate | Number of Patients Responding Weekly divided by the Number of Patients Assigned Weekly by provider |
| Patient (Micro) | Individuals Domain (e.g., roles, other personal attributes<sup>‡</sup>, | Age |  |
|  |  | Sex | Female or male |
|  |  | Race/Ethnicity | Non-Hispanic White, other, unknown or |
|  | and PROMs recipients) |  | declined to report |
|  |  | Education | Highest level of education attained; high school or less, college or above, unknown or declined to report |
|  |  | Insurance Type | Federal coverage (e.g., Medicare, Medicaid), commercial, other, and unknown or self-pay |
|  |  | Employment | Employed full-time, employed part-time, or student; unemployed, retired, or disabled; unknown |
|  |  | Preferred Language | English, non-English, or unknown |
|  |  | Comorbidity | NCI version of the Charlson comorbidity index |
|  |  | Cancer Type | Breast, prostate, lung, or other |
|  |  | Receipt of Surgery in the Last Six Months | Underwent an operation within the previous six months for any indication |
|  |  | Time Since Diagnosis | Number of months since the diagnosis of cancer |

CFIR: Consolidated Framework for Implementation Research. ‡: Elements were only available in the original CFIR (2009) but not the updated version in 2022.

